# Circulating metabolic biomarkers are consistently associated with incident type 2 diabetes in Asian and European populations – a metabolomics analysis in five prospective cohorts

**DOI:** 10.1101/2021.07.04.21259971

**Authors:** Jowy Yi Hoong Seah, Yueheng Hong, Anna Cichońska, Charumathi Sabanayagam, Simon Nusinovici, Tien Yin Wong, Ching-Yu Cheng, Pekka Jousilahti, Annamari Lundqvist, Markus Perola, Veikko Salomaa, E. Shyong Tai, Peter Würtz, Rob M. van Dam, Xueling Sim

**Affiliations:** Saw Swee Hock School of Public Health, National University of Singapore and National University Health System, Singapore; Singapore 117549, Singapore; Nightingale Health Ltd, Mannerheimintie 164a, 00300 Helsinki, Finland; Singapore Eye Research Institute, Singapore National Eye Centre, Singapore 169856, Singapore; Duke-NUS Medical School, Singapore 169857, Singapore; Yong Loo Lin School of Medicine, National University of Singapore and National University Health System, Singapore 117597, Singapore; Department of Public Health Solutions, Finnish Institute for Health and Welfare, P.O. Box 30, FI-00271 Helsinki, Finland; Research Program for Clinical and Molecular Metabolism, Faculty of Medicine, University of Helsinki, 00014 Helsinki, Finland; Departments of Exercise and Nutrition Sciences and Epidemiology, Milken Institute School of Public Health, The George Washington University, Washington, DC 20052

**Author notes:** These authors are co-corresponding senior authors., Rob M. van Dam. Department of Exercise and Nutrition Sciences, Milken Institute School of Public Health, The George Washington University, 950 New Hampshire Ave NW, Washington, DC 20052; Xueling Sim. Saw Swee Hock School of Public Health, 12 Science Drive 2, #10-01, Singapore 117549. These authors contributed equally (Jowy Yi Hoong Seah, Yueheng Hong, Anna Cichońska).

**Keywords:** NMR, metabolomics, type 2 diabetes, branched-chain amino acids, aromatic amino acids, glycoprotein acetyls, inflammation, fatty acids, lipoprotein, HDL, LDL, triglycerides, cholesterol

## Abstract

**Background:** While Asians have a higher risk of type 2 diabetes (T2D) than Europeans for a given BMI, it remains unclear whether the same markers of metabolic pathways are associated with diabetes.

**Objectives:** We evaluated associations between metabolic biomarkers and incident T2D in three major Asian ethnic groups (Chinese, Malay, and Indian) and a European population.

**Methods:** We analyzed data from adult males and females of two cohorts from Singapore (n = 6,393) consisting of Chinese, Malays and Indians, and three cohorts of European-origin participants from Finland (n = 14,558). We used nuclear magnetic resonance to quantify 154 circulating metabolic biomarkers at baseline and performed logistic regression to assess associations with T2D risk adjusted for age, sex, BMI and glycaemic markers.

**Results:** Of the 154 metabolic biomarkers, 59 were associated with higher risk of T2D in both Asians and Europeans (*P* < 0.0003; Bonferroni-corrected). These included branched-chain and aromatic amino acids, the inflammatory marker glycoprotein acetyls, total fatty acids, monounsaturated fatty acids, apolipoprotein B, larger very low-density lipoprotein particle sizes, and triglycerides. In addition, 13 metabolites were associated with a lower T2D risk in both populations including omega-6 polyunsaturated fatty acids and larger high-density lipoprotein particle sizes. Associations were consistent within the Asian ethnic groups (all *P*_het_ ≥ 0.05) and largely consistent for the Asian and European populations (*P*_het_ ≥ 0.05 for 128 of 154 metabolic biomarkers).

**Conclusion:** Metabolic biomarkers across several biological pathways were consistently associated with T2D risk in Asians and Europeans.

## Introduction

The global burden of type 2 diabetes (T2D) is high and rising [1, 2], with the prevalence expected to rise to 7079 cases per 100,000 in 2030, from 6059 cases per 100,000 in 2017 [2]. In randomized trials, pharmaceutical and lifestyle interventions substantially reduced the risk of T2D [3-5] in high-risk individuals. Variation in genetic, environmental and lifestyle exposures between individuals, coupled with heterogeneity in T2D [6, 7] has motivated omics biomarkers as intermediate phenotypes to T2D and the development of ‘precision medicine’ approaches to prevention [8-10]. Metabolic biomarkers reflect the dynamic and ongoing state of homeostasis and metabolism [11]. In recent years, the field of metabolomics has gained traction due to technological advances [12] and has uncovered various metabolites that are consistent predictors of T2D [13]. As many of these metabolic biomarkers are modifiable by diet or other lifestyle factors, information on metabolic risk factors may guide personalized interventions.

Previous studies on metabolic profiles and T2D risk were mostly conducted in populations of predominantly European ancestry, and it remains unclear if these findings can be generalized to different Asian ethnic groups. For example, individuals of Asian origin tend to develop T2D at a lower body mass index (BMI) and a younger age than those of European origin [7], and ethnic Indians are generally more insulin resistant than ethnic Malays or Chinese [14]. Although some metabolomics studies on T2D incidence have been conducted in East Asian (e.g. Chinese) populations, data on South East Asian (e.g. Malays) and South Asian (e.g. Indians) populations are limited [13].

We therefore evaluated whether associations between biomarkers reflecting a variety of biological pathways and diabetes incidence differed between Asian and European ethnic groups. We quantified amino acids, an inflammatory marker, fatty acids, lipoprotein and ketone bodies using nuclear magnetic resonance (NMR) metabolomic analysis in two multi-ethnic cohorts in Singapore and three cohorts in Finland. NMR metabolomics allows for highly reproducible identification and quantification of compounds [15, 16]. We subsequently performed association analyses of these metabolic measures with risk of T2D in three major Asian ethnic groups (Chinese, Malay, and Indian) and the European population.

## Methods

### Study Population

The Asian participants included in this study were from two population-based cohorts in Singapore that included ethnic Indians, Malays and Chinese: the Multiethnic Cohort (MEC) [17] study and the Singapore Epidemiology of Eye Diseases (SEED) cohort study [18, 19]. The European participants in our study were from three Finnish prospective population-based cohorts (FINRISK 2002, FINRISK 2012 and Health 2000) [20, 21]. All studies included questionnaires about socio-demographic characteristics and medical history, technician-measurement of anthropometric measures and blood pressure, and collection of blood samples. Details of the individual cohorts are provided in the **Supplemental Text**. Informed consent was obtained from all participants and ethical approvals were obtained from the respective ethics review boards.

For all cohorts, we excluded participants with diabetes, cardiovascular diseases or cancer at baseline. From the MEC cohort of 9180 participants, we identified 694 incident diabetes cases using the following criteria: (a) fasting plasma glucose ≥7.0 mmol/L or HbA1c ≥6.5 % (47.5 mmol/mol) based on the American Diabetes Association [22] or self-reported physician-diagnosed diabetes at follow-up, or (b) record linkage with a national database of medical diagnoses as part of public hospital, polyclinic, and subsidized GP visits). Using risk-set sampling, we selected 1315 controls matched for age (±5 years), sex, date of health screening (±2 years), and ethnicity from cohort participants who were alive, and did not have diabetes at the time the index case occurred. We used 6-year revisit data from 4384 SEED cohort participants among whom 391 developed diabetes during follow-up. Diabetes ascertainment at baseline and the revisit was based on random glucose ≥11.1 mmol/L or HbA1c ≥6.5 % (47.5 mmol/mol) [22], or a self-reported physician diagnosis. For the European cohorts, we used data on 14588 participants from FINRISK 2002, FINRISK 2012 and Health 2000, among whom 1055 developed diabetes during follow-up. Diabetes ascertainment at baseline was based on the National Social Insurance Institution Drug Purchase and Reimbursement Registries for purchases and reimbursements of purchases of hypoglycemic drugs, and the National Hospital Discharge Register for hospitalizations with diabetes as the diagnosis. Incident diabetes cases were identified by linking the participants to the registers described above with a unique personal identification code assigned to each Finnish citizen. Participants with prevalent or incident gestational diabetes were excluded.

### Metabolic Biomarker Quantification and Quality Control

Metabolic biomarkers were quantified from plasma (MEC, SEED Malays and European cohorts) and serum (SEED Chinese and SEED Indians) samples at baseline using a high-throughput ^1^H-NMR metabolomics platform developed by Nightingale Health Ltd. (Helsinki, Finland; nightingalehealth.com/; <u>biomarker quantification version 2016</u>). Details of the procedure have been described previously [23, 24]. The metabolic biomarkers quantified reflected several etiologically diverse pathways, such as amino acids, fatty acids, ketone bodies, and gluconeogenesis□lrelated measurements. In addition, lipoprotein measures including total concentration, total lipids, phospholipids, total cholesterol, cholesterol esters, free cholesterol and triglycerides within 14 lipoprotein subclasses were measured. 9 MEC and 20 SEED participants had >10% missing metabolic biomarker values and were excluded from subsequent analyses. For participants with metabolic biomarker values lower than detection level, we replaced values of 0 with a value equivalent to 0.9 multiplied by the non-zero minimum value of that measurement. Finally, we standardized the metabolic biomarker concentrations to z-scores, and all effect sizes were reported as per standard deviation increment to enable comparison of metabolic biomarker associations for measures with different units and concentration ranges. A total of 154 individual metabolic biomarkers were included in our analyses.

### Statistical analyses

We first calculated Pearson’s correlation coefficients for all 154 metabolic biomarkers. Logistic regression was then used to assess associations between circulating metabolic biomarkers with T2D. We did not use Cox regression because most of the T2D cases were detected during revisit and the time-to-event data therefore did not accurately reflect time from baseline to disease onset. For the matched MEC case-control study, conditional logistic regression model was used. In all studies, we adjusted for baseline age, sex, BMI, and fasting glucose (for MEC and Health 2000) or glycated hemoglobin (HbA1c, for non-fasting SEED, FINRISK 2002 and 2012 cohorts). For MEC, age^2^ was included in the conditional logistic model as there was evidence of non-linearity in the relationship between age and T2D risk. Logistic models were performed separately by study and ethnic group to account for differences between populations. Inverse variance-weighted fixed-effect meta-analysis was used to first pool ethnic-specific association statistics from the two Singapore cohorts, and then to combine association results from three ethnic groups. We also tested for heterogeneity using the Cochran Q heterogeneity. Bonferroni correction was used to account for multiple testing of 154 metabolic biomarkers and *P* <0.0003 (0.05/154) was considered statistically significant.

To test the joint effects of multiple metabolic biomarkers on risk of T2D, we first performed the variable selection as described below and subsequently included the selected metabolites in a multivariable logistic regression model. Due to the correlated nature of the markers, we considered pairwise correlations between the metabolic biomarkers from MEC (Supplement Table 1). For the set of T2D associated metabolic biomarkers in the association analysis above that considered a metabolic biomarker individually across Asian and European populations, we first sorted the metabolic biomarkers by their strength of association. Starting with the most significantly associated metabolic biomarker (index-associated metabolic biomarker), we iteratively excluded other associated metabolic biomarkers that had correlations ≥ 0.80 with the index-associated metabolic biomarker (correlation-pruning). The set of remaining metabolic biomarkers thus had a Bonferroni-adjusted significant association with T2D and pairwise correlations <0.80. Finally, we performed stepwise regression analysis with the criterion of *P*-value of 0.05 in a joint model that included the selected biomarkers in the Asian and European cohorts. All statistical analyses were conducted using R 3.3.2 (R Foundation for Statistical Computing, Vienna, Austria).

## Results

In the nested MEC case-control study, 694 incident T2D cases and 1315 matched controls were included in this study during a mean follow-up of 6.9 y (standard deviation SD: 2.4 y), and in the SEED cohort study, 391 incident T2D cases occurred in 4384 participants during an average of 6.2 y (SD: 1.1 y). In the European cohorts, 1055 incident T2D cases occurred in 14,558 participants during an average of 11.3 years (SD: 5.0 y). The average age was 54.0 y (SD: 9.8 y) for the Asian participants and 50.6 y (SD: 14.1 y) for the European participants, with similar representation of males and females for all populations, and Chinese, Malays and Indians for the Asian population (**Table 1**).

**Table 1.** Baseline characteristics of participants of the cohorts included in the study^1^

| Characteristic | MEC |  |  | SEED<sup>2</sup> |  |  | European cohorts |  |  |
| --- | --- | --- | --- | --- | --- | --- | --- | --- | --- |
|  | Chinese | Malay | Indian | Chinese | Malay | Indian | FINRISK 2002 | FINRISK 2012 | Health 2000 |
| No. cases / No. controls | 256/499 | 216/411 | 222/405 | 96/1825 | 132/1113 | 163/1055 | 350/3904 | 78/4475 | 627/6179 |
| Follow-up duration (y) | 7.4 ± 2.5 | 6.7 ± 2.5 | 6.6 ± 2.3 | 5.8 ± 0.9 | 7.1 ± 0.9 | 6.0 ± 0.9 | 13.8 ± 0.1 | 3.8 ± 0.1 | 15.1 ± 0.1 |
| Age at interview (y) | 53.6 ± 9.5 | 49.1 ± 9.9 | 47.8 ± 10.4 | 57 ± 8.6 | 54.8 ± 10 | 54.4 ± 8.4 | 48.5 ± 13.7 | 48.8 ± 13.8 | 53.2 ± 14.2 |
| Male [n (%)] | 356 (47.1) | 234 (37.3) | 257 (41) | 889 (46.3) | 563 (45.2) | 568 (46.6) | 1794 (45.9) | 2088 (46.7) | 2748 (44.5) |
| Education level<sup>3</sup> |  |  |  |  |  |  |  |  |  |
| Low | 243 (32.2) | 208 (33.2) | 199 (31.8) | 905 (47.1) | 773 (62.2) | 588 (48.4) | - | - | - |
| Intermediate | 415 (55) | 404 (64.5) | 356 (56.9) | 818 (42.6) | 455 (36.6) | 478 (39.3) | - | - | - |
| High | 97 (12.8) | 14 (2.2) | 71 (11.3) | 198 (10.3) | 15 (1.2) | 149 (12.3) | - | - | - |
| Body mass index (kg/m<sup>2</sup>) | 23.8 ± 3.7 | 26.9 ± 5 | 26.7 ± 4.9 | 23.4 ± 3.5 | 26 ± 4.7 | 25.7 ± 4.2 | 26.6 ± 4.5 | 26.6 ± 4.6 | 26.8 ± 4.6 |
| Fasting glucose (mmol/L) | 4.9 ± 0.6 | 5 ± 0.6 | 5.1 ± 0.6 | - | - | - | - | - | 5.4 ± 0.8 |
| HbA1c (%)<sup>4</sup> | 5.6 ± 0.4 | 5.6 ± 0.4 | 5.8 ± 0.4 | 5.8 ± 0.3 | 5.7 ± 0.4 | 5.7 ± 0.4 | 5.4 ± 0.4 | 5.2 ± 0.3 | - |
| Total cholesterol (mmol/L) | 5.3 ± 0.9 | 5.6 ± 1 | 5.3 ± 0.9 | 5.6 ± 1 | 5.7 ± 1.1 | 5.5 ± 1 | 5.6 ± 1.1 | 5.4 ± 1.0 | 6.0 ± 1.1 |
| HDL cholesterol (mmol/L) | 1.4 ± 0.3 | 1.3 ± 0.3 | 1.1 ± 0.3 | 1.4 ± 0.4 | 1.4 ± 0.3 | 1.1 ± 0.3 | 1.5 ± 0.4 | 1.5 ± 0.4 | 1.4 ± 0.4 |
| LDL cholesterol (mmol/L) | 3.4 ± 0.8 | 3.6 ± 0.9 | 3.5 ± 0.8 | 3.4 ± 0.9 | 3.6 ± 1 | 3.6 ± 0.8 | 3.4 ± 0.9 | 3.3 ± 0.9 | 3.8 ± 1.1 |
| Triglycerides (mmol/L) | 1.2 (0.9,1.7) | 1.2 (0.9,1.7) | 1.3 (0.9,1.8) | 1.4 (1.0,2.1) | 1.3 (0.7,2.1) | 1.6 (1.1,2.3) | 1.1 (0.8, 1.6) | 1.1 (0.8, 1.6) | 1.3 (1.0, 1.8) |
| DBP (mmHg) | 78.7 ± 11.3 | 76.4 ± 11.7 | 74.5 ± 11.5 | 77.6 ± 9.9 | 79.2 ± 11.0 | 77.9 ± 10.1 | 78.7 ± 11.4 | 81.3 ± 11.1 | 82.0 ± 11.0 |
| SBP (mmHg) | 134.2 ± 20.6 | 131.9 ± 20 | 125.3 ± 21.4 | 133.4 ± 18.4 | 141 ± 22.3 | 130.9 ± 18.7 | 135.0 ± 20.2 | 132.7 ± 18.6 | 134.2 ± 20.7 |

<sup>1</sup> Mean ± SD; median (Q1, Q3)
<sup>2</sup> SEED cohorts consist of a collection of three cohorts: Singapore Chinese Eye Study, Singapore Malay Eye Study and the Singapore Indian Eye Study. Participants of the SEED cohorts were not required to fast. LDL cholesterol was measured in the Singapore Chinese Eye Study and Singapore Indian Eye Study, and calculated in the Singapore Malay Eye Study.
<sup>3</sup> We classified education level as follows: low – none, Primary or equivalent, intermediate – Secondary, diploma, Junior College, high school or equivalent, and high – college degree and above. The data for European cohorts could not be accessed.
<sup>4</sup> Only a subset of participants in MEC (N = 598, 531, 516, for Chinese, Malay, and Indian) had HbA1c measurements. To convert HbA1c from % to mmol/mol, first multiply HbA1c (%) by 10.93 and then subtract 23.50.
Abbreviations: DBP, diastolic blood pressure, HbA1c, glycated hemoglobin A1c, HDL, high-density lipoprotein, LDL, low-density lipoprotein, MEC, Multiethnic Cohort, SEED, SBP, systolic blood pressure, Singapore Epidemiology of Eye Diseases, Q1, quantile 1, Q3, quantile 3

We observed high correlations among the branched-chain amino acids (BCAAs) leucine, isoleucine and valine, and concentrations of n-6 polyunsaturated (n-6 PUFA) fatty acids, PUFA, monounsaturated fatty acids (MUFA), saturated fatty acids and linoleic acid (18:2n-6) (all r>0.80) (**Supplemental Tables 1to 4**). In general, the lipid components (total lipids, phospholipids, cholesterol esters, free cholesterol) with the exception of triglycerides within a particular lipoprotein subclass were also highly correlated. Overall, 59 of the 154 metabolic biomarkers were consistently associated with a higher risk and 13 with a lower risk of T2D in both Asians and Europeans (*P*_*meta*_ < 0.0003; Bonferroni-corrected; *P*_het_ < 0.05 for 12 metabolic biomarkers) (**Table 2, Figure 1** and **Supplemental Table 5)** after adjustment for age, sex, BMI and either fasting glucose or HbA1c. For the population-specific meta-analysis, we observed little heterogeneity in association across the three Asian ethnic groups (*P*_het_ ≥ 0.05 for all 154 metabolic biomarkers) and across the three European cohorts (*P*_het_ ≥ for 144 of 154 metabolic biomarkers). Of the 29 metabolic biomarkers for which we did observe evidence for heterogeneity between the Asian and European populations, 23 were lipoprotein measures.

**Table 2.**
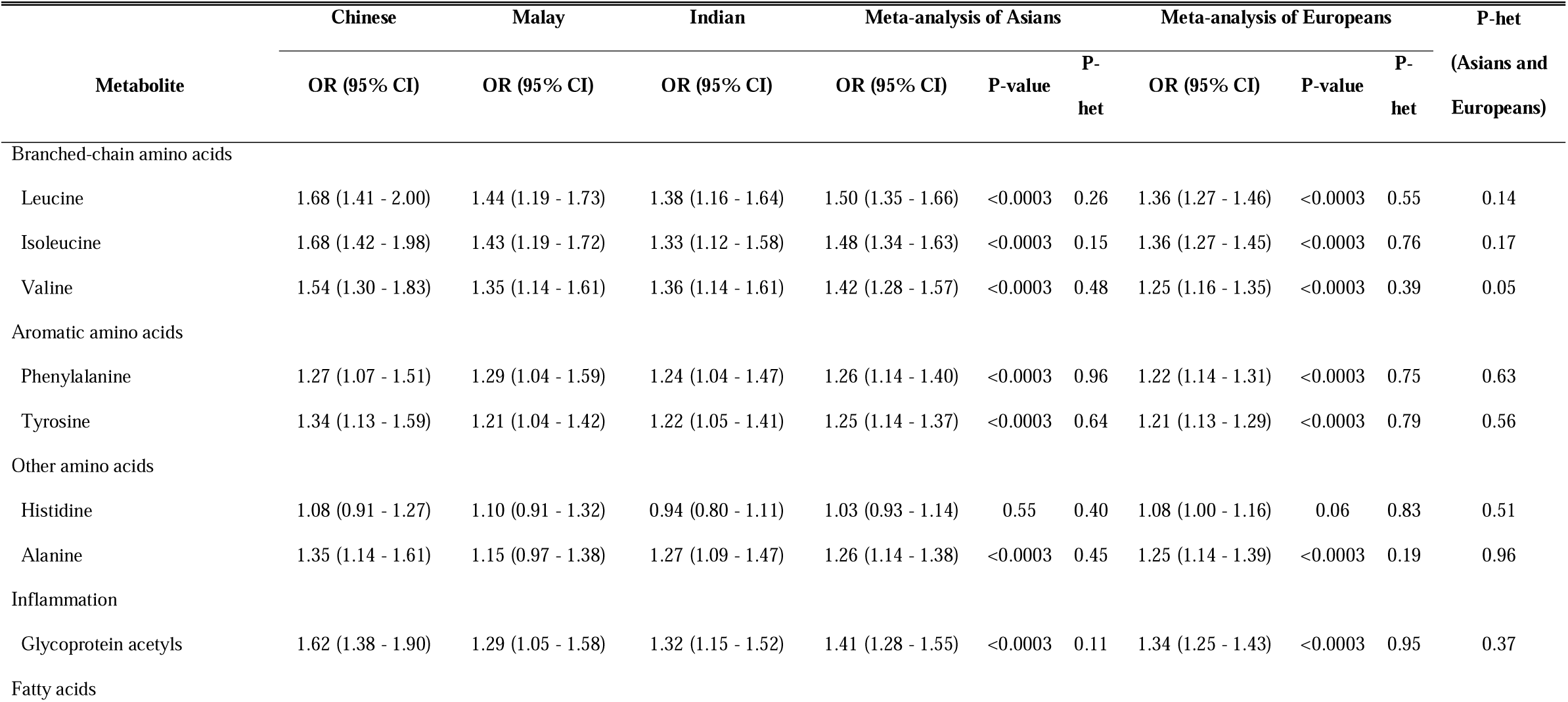
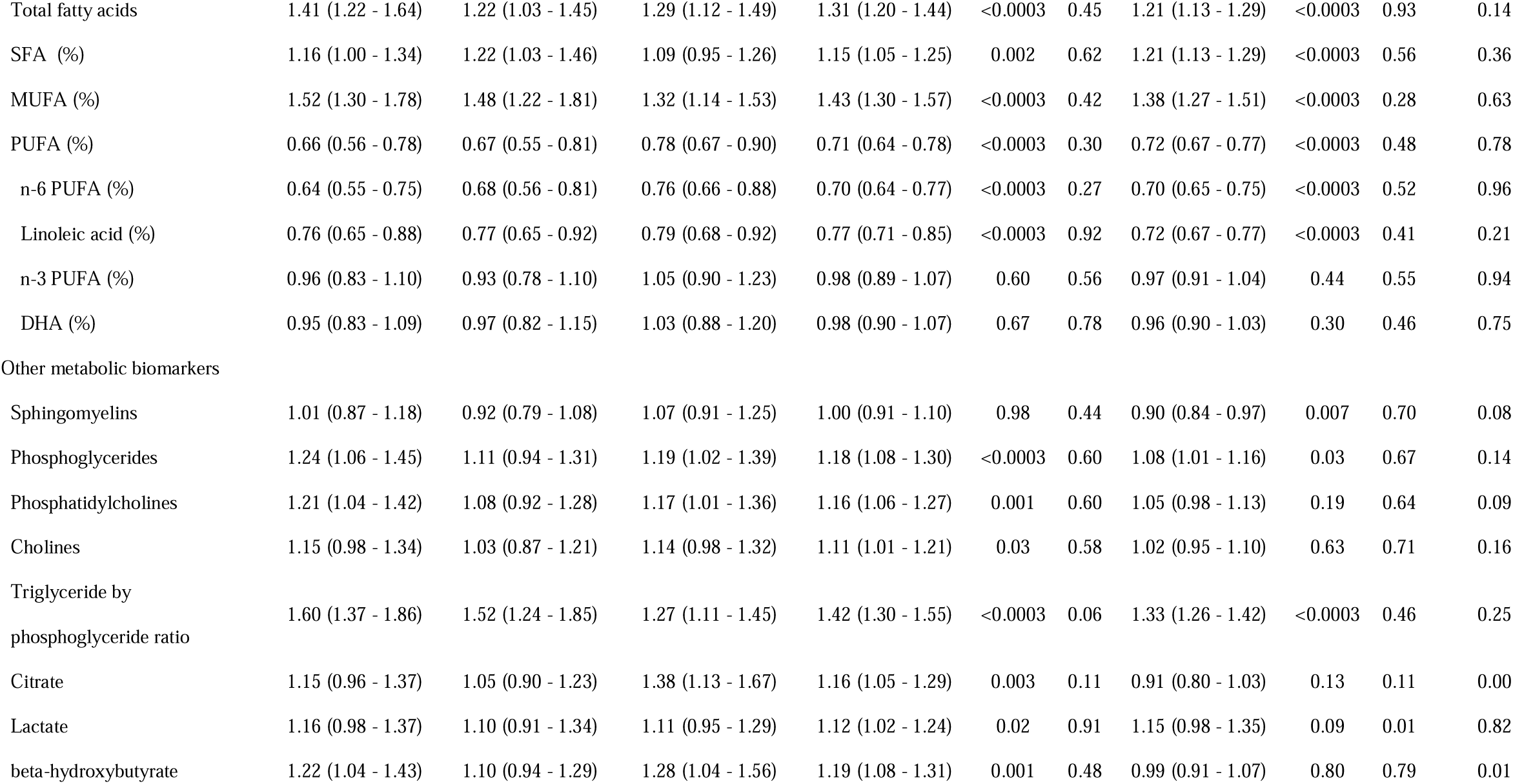
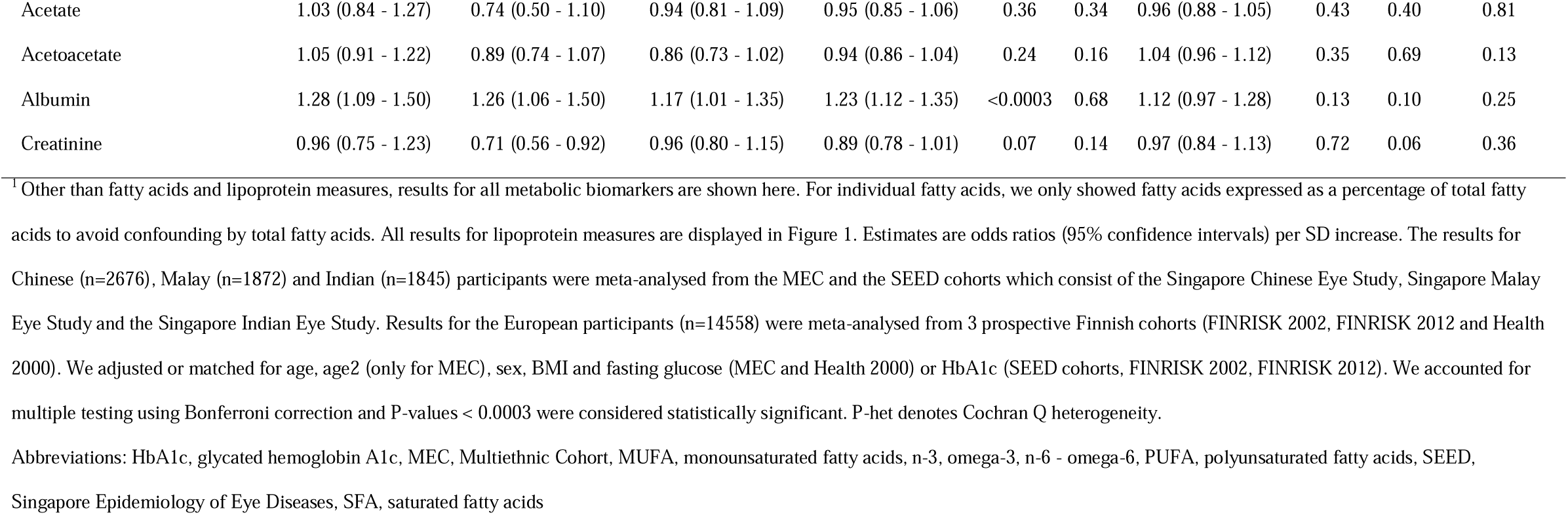
Associations between non-lipoprotein metabolic biomarkers and type 2 diabetes^1^

| Metabolite | Chinese | Malay | Indian | Meta-analysis of Asians |  |  | Meta-analysis of Europeans |  |  | P-het |
| --- | --- | --- | --- | --- | --- | --- | --- | --- | --- | --- |
|  | OR (95% CI) | OR (95% CI) | OR (95% CI) | OR (95% CI) | P-value | P-het | OR (95% CI) | P-value | P-het | (Asians and Europeans) |
| Branched-chain amino acids |  |  |  |  |  |  |  |  |  |  |
| Leucine | 1.68 (1.41 - 2.00) | 1.44 (1.19 - 1.73) | 1.38 (1.16 - 1.64) | 1.50 (1.35 - 1.66) | <0.0003 | 0.26 | 1.36 (1.27 - 1.46) | <0.0003 | 0.55 | 0.14 |
| Isoleucine | 1.68 (1.42 - 1.98) | 1.43 (1.19 - 1.72) | 1.33 (1.12 - 1.58) | 1.48 (1.34 - 1.63) | <0.0003 | 0.15 | 1.36 (1.27 - 1.45) | <0.0003 | 0.76 | 0.17 |
| Valine | 1.54 (1.30 - 1.83) | 1.35 (1.14 - 1.61) | 1.36 (1.14 - 1.61) | 1.42 (1.28 - 1.57) | <0.0003 | 0.48 | 1.25 (1.16 - 1.35) | <0.0003 | 0.39 | 0.05 |
| Aromatic amino acids |  |  |  |  |  |  |  |  |  |  |
| Phenylalanine | 1.27 (1.07 - 1.51) | 1.29 (1.04 - 1.59) | 1.24 (1.04 - 1.47) | 1.26 (1.14 - 1.40) | <0.0003 | 0.96 | 1.22 (1.14 - 1.31) | <0.0003 | 0.75 | 0.63 |
| Tyrosine | 1.34 (1.13 - 1.59) | 1.21 (1.04 - 1.42) | 1.22 (1.05 - 1.41) | 1.25 (1.14 - 1.37) | <0.0003 | 0.64 | 1.21 (1.13 - 1.29) | <0.0003 | 0.79 | 0.56 |
| Other amino acids |  |  |  |  |  |  |  |  |  |  |
| Histidine | 1.08 (0.91 - 1.27) | 1.10 (0.91 - 1.32) | 0.94 (0.80 - 1.11) | 1.03 (0.93 - 1.14) | 0.55 | 0.40 | 1.08 (1.00 - 1.16) | 0.06 | 0.83 | 0.51 |
| Alanine | 1.35 (1.14 - 1.61) | 1.15 (0.97 - 1.38) | 1.27 (1.09 - 1.47) | 1.26 (1.14 - 1.38) | <0.0003 | 0.45 | 1.25 (1.14 - 1.39) | <0.0003 | 0.19 | 0.96 |
| Inflammation |  |  |  |  |  |  |  |  |  |  |
| Glycoprotein acetyls | 1.62 (1.38 - 1.90) | 1.29 (1.05 - 1.58) | 1.32 (1.15 - 1.52) | 1.41 (1.28 - 1.55) | <0.0003 | 0.11 | 1.34 (1.25 - 1.43) | <0.0003 | 0.95 | 0.37 |
| Fatty acids |  |  |  |  |  |  |  |  |  |  |
| Total fatty acids | 1.41 (1.22 - 1.64) | 1.22 (1.03 - 1.45) | 1.29 (1.12 - 1.49) | 1.31 (1.20 - 1.44) | <0.0003 | 0.45 | 1.21 (1.13 - 1.29) | <0.0003 | 0.93 | 0.14 |
| SFA (%) | 1.16 (1.00 - 1.34) | 1.22 (1.03 - 1.46) | 1.09 (0.95 - 1.26) | 1.15 (1.05 - 1.25) | 0.002 | 0.62 | 1.21 (1.13 - 1.29) | <0.0003 | 0.56 | 0.36 |
| MUFA (%) | 1.52 (1.30 - 1.78) | 1.48 (1.22 - 1.81) | 1.32 (1.14 - 1.53) | 1.43 (1.30 - 1.57) | <0.0003 | 0.42 | 1.38 (1.27 - 1.51) | <0.0003 | 0.28 | 0.63 |
| PUFA (%) | 0.66 (0.56 - 0.78) | 0.67 (0.55 - 0.81) | 0.78 (0.67 - 0.90) | 0.71 (0.64 - 0.78) | <0.0003 | 0.30 | 0.72 (0.67 - 0.77) | <0.0003 | 0.48 | 0.78 |
| n-6 PUFA (%) | 0.64 (0.55 - 0.75) | 0.68 (0.56 - 0.81) | 0.76 (0.66 - 0.88) | 0.70 (0.64 - 0.77) | <0.0003 | 0.27 | 0.70 (0.65 - 0.75) | <0.0003 | 0.52 | 0.96 |
| Linoleic acid (%) | 0.76 (0.65 - 0.88) | 0.77 (0.65 - 0.92) | 0.79 (0.68 - 0.92) | 0.77 (0.71 - 0.85) | <0.0003 | 0.92 | 0.72 (0.67 - 0.77) | <0.0003 | 0.41 | 0.21 |
| n-3 PUFA (%) | 0.96 (0.83 - 1.10) | 0.93 (0.78 - 1.10) | 1.05 (0.90 - 1.23) | 0.98 (0.89 - 1.07) | 0.60 | 0.56 | 0.97 (0.91 - 1.04) | 0.44 | 0.55 | 0.94 |
| DHA (%) | 0.95 (0.83 - 1.09) | 0.97 (0.82 - 1.15) | 1.03 (0.88 - 1.20) | 0.98 (0.90 - 1.07) | 0.67 | 0.78 | 0.96 (0.90 - 1.03) | 0.30 | 0.46 | 0.75 |
| Other metabolic biomarkers |  |  |  |  |  |  |  |  |  |  |
| Sphingomyelins | 1.01 (0.87 - 1.18) | 0.92 (0.79 - 1.08) | 1.07 (0.91 - 1.25) | 1.00 (0.91 - 1.10) | 0.98 | 0.44 | 0.90 (0.84 - 0.97) | 0.007 | 0.70 | 0.08 |
| Phosphoglycerides | 1.24 (1.06 - 1.45) | 1.11 (0.94 - 1.31) | 1.19 (1.02 - 1.39) | 1.18 (1.08 - 1.30) | <0.0003 | 0.60 | 1.08 (1.01 - 1.16) | 0.03 | 0.67 | 0.14 |
| Phosphatidylcholines | 1.21 (1.04 - 1.42) | 1.08 (0.92 - 1.28) | 1.17 (1.01 - 1.36) | 1.16 (1.06 - 1.27) | 0.001 | 0.60 | 1.05 (0.98 - 1.13) | 0.19 | 0.64 | 0.09 |
| Cholines | 1.15 (0.98 - 1.34) | 1.03 (0.87 - 1.21) | 1.14 (0.98 - 1.32) | 1.11 (1.01 - 1.21) | 0.03 | 0.58 | 1.02 (0.95 - 1.10) | 0.63 | 0.71 | 0.16 |
| Triglyceride by<br>phosphoglyceride ratio | 1.60 (1.37 - 1.86) | 1.52 (1.24 - 1.85) | 1.27 (1.11 - 1.45) | 1.42 (1.30 - 1.55) | <0.0003 | 0.06 | 1.33 (1.26 - 1.42) | <0.0003 | 0.46 | 0.25 |
| Citrate | 1.15 (0.96 - 1.37) | 1.05 (0.90 - 1.23) | 1.38 (1.13 - 1.67) | 1.16 (1.05 - 1.29) | 0.003 | 0.11 | 0.91 (0.80 - 1.03) | 0.13 | 0.11 | 0.00 |
| Lactate | 1.16 (0.98 - 1.37) | 1.10 (0.91 - 1.34) | 1.11 (0.95 - 1.29) | 1.12 (1.02 - 1.24) | 0.02 | 0.91 | 1.15 (0.98 - 1.35) | 0.09 | 0.01 | 0.82 |
| beta-hydroxybutyrate | 1.22 (1.04 - 1.43) | 1.10 (0.94 - 1.29) | 1.28 (1.04 - 1.56) | 1.19 (1.08 - 1.31) | 0.001 | 0.48 | 0.99 (0.91 - 1.07) | 0.80 | 0.79 | 0.01 |
| Acetate | 1.03 (0.84 - 1.27) | 0.74 (0.50 - 1.10) | 0.94 (0.81 - 1.09) | 0.95 (0.85 - 1.06) | 0.36 | 0.34 | 0.96 (0.88 - 1.05) | 0.43 | 0.40 | 0.81 |
| Acetoacetate | 1.05 (0.91 - 1.22) | 0.89 (0.74 - 1.07) | 0.86 (0.73 - 1.02) | 0.94 (0.86 - 1.04) | 0.24 | 0.16 | 1.04 (0.96 - 1.12) | 0.35 | 0.69 | 0.13 |
| Albumin | 1.28 (1.09 - 1.50) | 1.26 (1.06 - 1.50) | 1.17 (1.01 - 1.35) | 1.23 (1.12 - 1.35) | <0.0003 | 0.68 | 1.12 (0.97 - 1.28) | 0.13 | 0.10 | 0.25 |
| Creatinine | 0.96 (0.75 - 1.23) | 0.71 (0.56 - 0.92) | 0.96 (0.80 - 1.15) | 0.89 (0.78 - 1.01) | 0.07 | 0.14 | 0.97 (0.84 - 1.13) | 0.72 | 0.06 | 0.36 |

<sup>†</sup>Other than fatty acids and lipoprotein measures, results for all metabolic biomarkers are shown here. For individual fatty acids, we only showed fatty acids expressed as a percentage of total fatty acids to avoid confounding by total fatty acids. All results for lipoprotein measures are displayed in Figure 1. Estimates are odds ratios (95% confidence intervals) per SD increase. The results for Chinese (n=2676), Malay (n=1872) and Indian (n=1845) participants were meta-analysed from the MEC and the SEED cohorts which consist of the Singapore Chinese Eye Study, Singapore Malay Eye Study and the Singapore Indian Eye Study. Results for the European participants (n=14558) were meta-analysed from 3 prospective Finnish cohorts (FINRISK 2002, FINRISK 2012 and Health 2000). We adjusted or matched for age, age2 (only for MEC), sex, BMI and fasting glucose (MEC and Health 2000) or HbA1c (SEED cohorts, FINRISK 2002, FINRISK 2012). We accounted for multiple testing using Bonferroni correction and P-values < 0.0003 were considered statistically significant. P-het denotes Cochran Q heterogeneity.
Abbreviations: HbA1c, glycated hemoglobin A1c, MEC, Multiethnic Cohort, MUFA, monounsaturated fatty acids, n-3, omega-3, n-6 - omega-6, PUFA, polyunsaturated fatty acids, SEED, Singapore Epidemiology of Eye Diseases, SFA, saturated fatty acids

**Figure 1.**
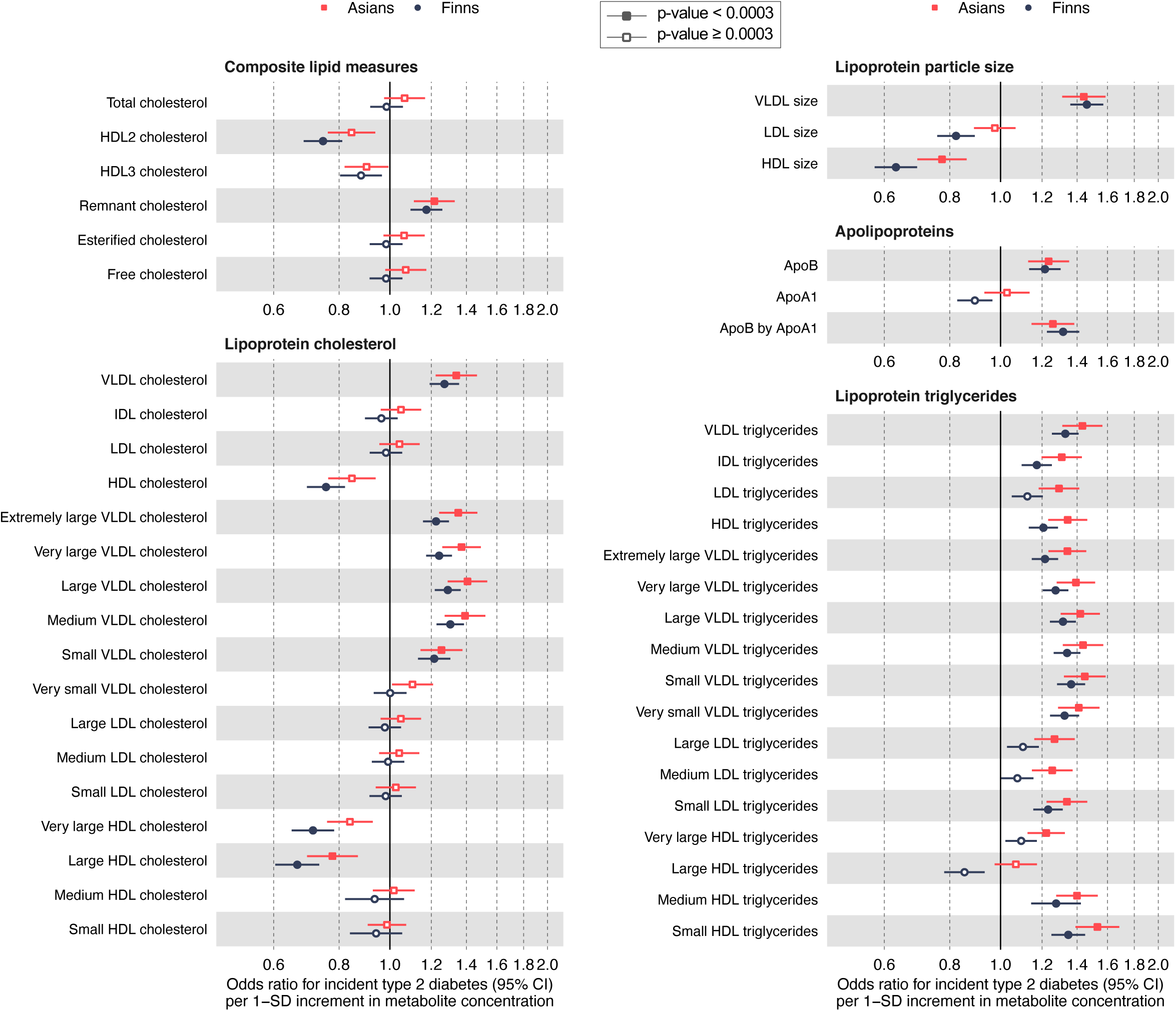
Associations between baseline circulating lipoprotein measures and risk of incident type 2 diabetes. Values are ORs (95% CIs) per 1 SD increment in metabolic biomarkers levels. The results for Asian participants (n=6393) were meta-analysed using data from Chinese (n=2676), Malay (n=1872) and Indian (n=1845) participants of the MEC and the SEED cohorts which consist of the Singapore Chinese Eye Study, Singapore Malay Eye Study and the Singapore Indian Eye Study. Results for the European participants (n=14558) were meta-analysed from 3 prospective Finnish cohorts (FINRISK 2002, FINRISK 2012 and Health 2000). We adjusted or matched for age, age^2^ (only for MEC), sex, BMI and fasting glucose (MEC and Health 2000) or HbA1c (SEED cohorts, FINRISK 2002 and FINRISK 2012). We accounted for multiple testing using Bonferroni correction and *P*-values < 0.0003 were considered statistically significant. Abbreviations: ApoA1, apolipoprotein A1, ApoB, apolipoprotein B, HbA1c, glycated hemoglobin A1c, HDL, high-density lipoprotein, IDL, intermediate-density lipoprotein, LDL, low-density lipoprotein, MEC, Multiethnic Cohort, SEED, Singapore Epidemiology of Eye Diseases, VLDL, very low-density lipoprotein

### Amino Acids and Inflammation

Higher concentrations of BCAAs (leucine, isoleucine, and valine), aromatic amino acids (phenylalanine and tyrosine), and alanine were significantly and consistently associated with higher T2D risk in both Asians and Europeans (all *P*_het_ > 0.05). The strongest association was observed for leucine (Asians: OR 1.50 per 1 SD increment, 95% CI 1.35 - 1.66; Europeans: OR 1.36, 95% CI 1.27 - 1.46). The inflammatory marker glycoprotein acetyls (GlycA) was also directly associated with risk of T2D (Asians: OR 1.41, 95% CI 1.28 - 1.55; Europeans: OR 1.34, 95% CI 1.25 - 1.43; *P*_het_ = 0.37). GlycA remained directly associated with T2D risk after further adjustments for CRP (data available only for MEC and the European cohorts, Asians: OR 1.94, 95% CI 1.63 - 2.31; Europeans: OR 1.34, 95% CI 1.25 - 1.44).

### Fatty Acids

Higher total concentrations of fatty acids were significantly associated with higher T2D risk. Among the different types of fatty acids, a higher proportion of MUFA was consistently associated with a higher T2D risk (Asians: OR 1.43, 95% CI 1.30 - 1.57; Europeans: OR 1.38, 95% CI 1.27 - 1.51), and a higher proportion of n-6 PUFA with a lower risk (Asians: OR 0.70, 95% CI 0.64 - 0.77; Europeans: OR 0.70, 95% CI 0.65 - 0.75). The proportion of saturated fat (SFA) was directly associated with T2D risk, but this was not statistically significant after Bonferroni correction in Asians (Asians: OR 1.15, 95% CI: 1.05 - 1.25; Europeans: OR 1.21, 95% CI 1.13 - 1.29). n-3 PUFAs were not significantly associated with risk of T2D in any ethnic group. There was no evidence of heterogeneity for associations of fatty acids with T2D between the Asian and European populations (all *P*_het_ > 0.05).

### Lipoprotein Measures

We evaluated lipoprotein particle sizes, apolipoproteins and various lipoprotein measures including total concentrations, total lipids, phospholipids, total cholesterol, cholesterol esters, free cholesterol and triglycerides for 14 lipoprotein subclasses, in relation to T2D (**Figure 1** and **Supplemental Table 5**).

Higher levels of apolipoprotein B were associated with higher T2D risk in both Asians (OR 1.24, 95% CI 1.13 - 1.35) and Europeans (OR 1.22, 95% CI 1.13 - 1.30; *P*_het_ = 0.45). Larger VLDL particle size was among the metabolic biomarkers with the strongest direct associations with T2D risk (Asians: OR 1.44, 95% CI 1.31 - 1.59; Europeans: OR 1.46, 95% CI 1.36 - 1.57; *P*_het_ = 0.83). Larger LDL particle sizes were inversely associated with incident T2D risk in Europeans (OR 0.82, 95% CI 0.76 - 0.89), but not in Asians (Asians: OR 0.98, 95% CI 0.89 - 1.07; *P*_het_ = 0.01). Larger HDL particle sizes were also more strongly inversely associated with T2D risk in Europeans (OR 0.63, 95% CI 0.58 - 0.69) as compared with Asians (OR 0.77, 95% CI 0.69 - 0.86; *P*_het_ = 0.01).

The concentration of cholesterol in very low-density lipoprotein (VLDL) particles was generally associated with higher risk of T2D in both Asians and Europeans (all *P*_het_ > 0.05), with the strongest association observed for cholesterol in large VLDL particles (Asians: OR 1.41, 95% CI 1.29 - 1.53; Europeans: OR 1.29, 95% CI 1.22 - 1.37; *P*_het_ = 0.10). Because the lipid components (total lipids, phospholipids, cholesterol esters, free cholesterol, triglycerides) in VLDL particles were highly correlated (**Supplemental Tables 1 to 4**), particularly in extremely large VLDL, very large VLDL, large VLDL and medium VLDL (Pearson’s r>0.85), these lipid components were similarly associated with T2D risk as VLDL cholesterol.

Total high-density lipoprotein (HDL) cholesterol was inversely associated with T2D risk in Asians (OR 0.85, 95% CI 0.76 - 0.94) and Europeans (OR 0.76, 95% CI 0.69 - 0.82; *P*_het_ = 0.09), although this did not reach Bonferroni-corrected significance in Asians. These inverse associations appeared to be driven by cholesterol in very large and large HDL particles, which were stronger in Europeans than in Asians (very large HDL: *P*_het_=0.02; large HDL: *P*_het_=0.04). Other lipid components of HDL (total lipids, phospholipids, cholesterol esters, free cholesterol) with the exception of triglycerides were similarly associated with T2D risk as the cholesterol component of HDL. There were no significant associations between intermediate-density lipoprotein (IDL) cholesterol and low-density lipoprotein (LDL) cholesterol concentrations and T2D risk.

Elevated concentrations of total triglycerides were significantly associated with a higher risk of T2D in Asians (OR 1.43, 95% 1.31 - 1.57) and Europeans (OR 1.32, 95% CI 1.25 - 1.40; *P*_het_ = 0.14). Results were similar when we analyzed triglycerides in subclasses of VLDL, LDL, and HDL, with the exception of triglycerides in large HDL.

### Other Metabolic Biomarkers

We also evaluated ketone bodies, glycerides, phospholipids, and metabolites related to glycolysis and fluid balance (**Table 2**). Higher phosphoglyceride concentrations were associated with T2D risk but this remained significant after Bonferroni correction only in Asians (OR 1.18, 95% CI 1.08 - 1.30) and not in Europeans (OR 1.08, 95% CI 1.01, 1.16; *P*_het_ = 0.14). Similarly, higher albumin concentrations were significantly associated with a higher risk of T2D in Asians (OR 1.23, 95% CI: 1.12 - 1.35), but not Europeans (OR 1.12, 95% CI 0.97 - 1.28; *P*_het_ = 0.25).

### Joint Effects of Metabolic Biomarkers on Risk of T2D

Using the correlation-pruning method, 12 metabolic biomarkers in MEC were significantly associated with T2D with pairwise correlations with all other biomarkers <0.80. These were isoleucine, valine, phenylalanine, tyrosine, alanine, GlycA, unsaturation index, proportion of n-6 PUFA, proportion of MUFA, VLDL size, small VLDL particle concentration, and very large HDL phospholipids. Stepwise regression selected a multivariable model that simultaneously included isoleucine, GlycA, unsaturation index, and very large HDL phospholipids (**Supplemental Table 6**). Higher levels of isoleucine (Asians: OR 1.25, 95% CI 1.10 - 1.42; Europeans: 1.18, 95% CI 1.06 - 1.31) and GlycA (Asians: OR 1.17, 95% CI 1.03 - 1.33; Europeans: OR 1.06, 95% CI 0.95 - 1.18) were associated with higher T2D risk, and very large HDL phospholipids with lower T2D risk (Asians: OR 0.88, 95% CI 0.77 - 0.99; Europeans: OR 0.70, 95% CI 0.62 - 0.78). There was no independent significant association between unsaturation index and T2D risk. We observed evidence of heterogeneity between the Asian and European populations only for very large HDL phospholipids (*P*_het_ = 0.01).

## Discussion

We quantified 154 metabolic biomarkers using NMR technology in plasma and serum samples from five prospective cohort studies representing three major ethnic groups in Asia (Chinese, Malay, and Indian) and a European population. While there have been previous metabolomics studies in Asians and individuals of European descent, these were mostly conducted separately in ethnically homogenous populations with cross-sectional study design, different metabolomics platforms or statistical methods that did not facilitate direct comparison of results [13, 25]. Here, we report that branched-chain amino acids, aromatic amino acids, alanine, the inflammatory marker GlycA, total fatty acids, the proportion of MUFA, apolipoprotein B, larger VLDL particle sizes and triglycerides were consistently associated with a higher T2D risk in Asians and Europeans. Furthermore, the proportion of n-6 PUFA and larger HDL particle sizes were consistently associated with a lower T2D risk in Asians and Europeans. Overall, associations were consistent across the Asian ethnic groups, and largely consistent for the Asian and European populations.

Our finding that higher concentrations of BCAAs (leucine, isoleucine and valine) were associated with a higher risk of T2D is consistent with the results of previous prospective studies [13, 23, 25-30]. Results from Mendelian randomization studies suggest that BCAA metabolites play a causal role in the pathogenesis of T2D [30]. In line with our results, the aromatic amino acids phenylalanine and tyrosine were directly associated with T2D risk in a meta-analysis of prospective studies [13], and alanine was directly associated with T2D risk in a Japanese cohort [27]. Elevated concentrations of aromatic amino acids and alanine have been hypothesized to induce insulin resistance, possibly by inhibition of glucose transport and phosphorylation in skeletal muscle [13, 31, 32].

A higher concentration of the inflammatory marker GlycA was associated with a higher risk of T2D in our study, which is compatible with results from the Dutch PREVEND study and the U.S. Women’s Health Study [33, 34]. GlycA is a composite NMR measure that arises from the *N*-acetyl methyl group protons of oligosaccharide moieties of acute-phase proteins [35], proteins whose concentrations change in response to inflammation. As opposed to CRP, which is characterized by multi-fold elevation in acute state, GlycA levels fluctuate less on short-term bases and may therefore capture different aspects of the inflammatory response [36], a hypothesis supported by our results and results of the PREVEND study as GlycA remains directly associated with T2D risk after adjusting for CRP [33]. However, in the Women’s Health Study, the association between GlycA and incident T2D was significantly attenuated after adjusting for CRP [34]. It is plausible that low-grade inflammation is involved in the pathogenesis of T2D through insulin resistance and beta-cell dysfunction [37, 38].

Our report of a direct association between circulating MUFA and T2D risk is consistent with a previous finding in a Finnish cohort [39], but not with the Atherosclerosis Risk in Communities (ARIC) study [40]. Because MUFA can be synthesized endogenously from SFA [41], MUFAs concentrations may reflect SFA intake. We also observed a direct association between circulating SFA and T2D risk although this was not significant in Asians. In the ARIC study [40] and a Finnish cohort [39], a higher proportion of SFA was also associated with higher T2D risk. It has been hypothesized that high SFA concentrations promote insulin resistance and are lipotoxic to beta-cells [42, 43]. Our study further showed that higher proportions of n-6 PUFAs were associated with a lower risk of T2D, concordant with a previous meta-analysis of prospective cohort studies [44]. The association may be partly mediated by increasing insulin sensitivity through increasing cellular membrane fluidity [45, 46], particularly of skeletal muscle cells and hepatocytes [45, 46], and by acting as ligands for peroxisome proliferator-activated receptor gamma [47, 48]. These hypotheses are supported by a meta-analysis of randomized feeding trials that showed improvements in glucose-insulin homeostasis for dietary PUFA compared to SFA or MUFA [49]. In a recent Mendelian randomization study, genetic predisposition to higher levels of linoleic acid was associated with a lower risk of T2D [50], supporting a causal effect of linoleic acid on T2D. The lack of association between the relative fraction of n-3 PUFA and T2D risk is consistent with a previous meta-analysis of prospective studies [51].

We also identified several patterns in the lipoprotein profile that were consistently associated with T2D risk across ethnic groups. Larger VLDL particles were directly associated with T2D risk, similar to existing evidence from prospective studies [23, 52-54]. VLDL is mostly comprised of triglycerides, and larger particles carry more triglycerides [54]. The causal relationship between circulating triglycerides and T2D is still a matter of debate [55] and there was no consensus among studies on genetically influenced triglyceride levels and T2D incidence [56, 57]. Our finding that the concentration of larger HDL particles was inversely associated with T2D risk also agrees with previous prospective studies in western populations [23, 52-54]. While HDL lipid components (e.g. phospholipids, cholesterol esters, free cholesterol) in very large and large HDL particles were associated with lower risk of T2D, they were not associated with T2D risk in medium and small HDL particles. This suggests that the inverse associations observed for HDL lipid components were driven by particle size rather than lipid composition. Larger HDL particles have been hypothesized to be more efficient in cholesterol efflux capacity than smaller HDL particles [58, 59]. HDL particles may also have other anti-diabetic properties such as promoting insulin secretion and glucose uptake in skeletal muscle cells [60].

Our study’s strengths included the use of large population-based cohorts collected in diverse populations with reasonable follow-up times that allowed us to have sufficient statistical power to evaluate the associations separately in each ethnic group and for heterogeneity between populations. However, we also acknowledge certain limitations. First, a limitation of NMR technology compared to mass spectrometry is its lower sensitivity, and we could not quantify metabolic biomarkers that were below the detection limit of NMR. Second, participants of the SEED cohorts were not required to fast and this may have affected levels of certain metabolic biomarkers such as triglycerides and amino acids [61, 62]. However, this would have more likely weakened than strengthened the observed associations. Third, different T2D case ascertainments were used for the cohorts and this may have affected comparability of the results. However, despite different methodologies in case ascertainments, we obtained consistent results across the different cohorts. Finally, we cannot rule out residual confounding due to unmeasured or imperfectly measured risk factors.

Our results may have implications in clinical practice, including the prevention and treatment, and early detection and prognosis of T2D. The metabolic biomarkers associated with T2D risk in our study are modifiable and may thus provide targets for lifestyle and pharmacological interventions for prevention and treatment. For example, replacing dietary SFA with PUFA has long been part of dietary recommendation to reduce cardiovascular disease risk [63] and may also reduce T2D risk by improving plasma fatty acid profiles. Similarly, dietary BCAAs and aromatic amino acids are primarily from animal products, such as meat, and lower consumption may reduce T2D risk partly by improving circulating amino acids [64-66] and insulin sensitivity [67]. Furthermore, cholesteryl ester transfer protein inhibitors are a drug class that is known to preferentially increase larger HDL particles [68, 69]. In a meta-analysis of four randomized trials, these drugs significantly reduced T2D risk although it is unclear if this effect is caused solely by changes in circulating HDL [68-70]. Because these metabolic biomarkers reflect modifiable pathways, metabolic profiling data could inform ‘precision medicine’ approaches that prioritize specific preventive interventions for individuals at high risk of T2D. Various biomarkers such as BCAAs and aromatic amino acids may be etiologically involved in insulin resistance [31, 71], and the inclusion of these biomarkers to prediction models of T2D improved area under curve (AUC) [72]. As the biological pathways involved in the pathogenesis of T2D appear to be similar in Asians and Europeans, these interventions and approaches for detection and prediction may be effective for both populations.

Several physiological differences have been proposed to contribute to the higher susceptibility to developing T2D in Asians than Europeans [7]. For example, greater visceral adiposity and less lean body mass may contribute to the higher rates of T2D in South Asians as compared with European ancestry populations [7, 73, 74]. The generally consistent associations for metabolic biomarkers across diverse ethnic groups in our study suggest that differences in the degree of metabolic disturbances in the evaluated pathways rather than differences in the pathways involved contribute to ethnic differences in risk of T2D. However, there may be exceptions for selected biological pathways. For example, South Asians have been found to have smaller mean HDL particle sizes as compared with European ancestry populations [75] and this may contribute to the higher risk of T2D in South Asians through less efficient reverse cholesterol transport and dyslipidemia [58]. Our observation of possibly stronger inverse associations between HDL and T2D risk in Europeans than Asians, particularly for very large HDL, is novel and warrants further studies.

We found consistent associations between metabolic signatures and risk of T2D in prospective cohorts in Asia and Europe. Metabolic aberrations that spanned multiple pathways, including amino acids, inflammation, fatty acids and lipoproteins were similarly associated with T2D risk in major Asian ethnic groups and Europeans despite environmental, lifestyle, and physiological differences. Our results suggest that these are candidates for better prediction of T2D in different ethnic groups that require further evaluation in prognostic studies. Moreover, these metabolic biomarkers are modifiable and may be targets for preventive interventions across different ethnic groups. Metabolic profiles may inform future personalized interventions for the prevention of T2D in increasingly cosmopolitan populations.

## Data Availability

Data described in the manuscript, code book and analytic code will be made available upon request pending application and approval.

## Abbreviations

ARIC: Atherosclerosis Risk in Communities
BCAA: branched-chain amino acids
BMI: body mass index
GlycA: glycoprotein acetyls
HbA1c: glycated hemoglobin
HDL: high-density lipoprotein
IDL: intermediate-density lipoprotein
LDL: low-density lipoprotein
MEC: Multiethnic Cohort
MUFA: monounsaturated fat
NMR: nuclear magnetic resonance
PUFA: polyunsaturated fat
SEED: Singapore Epidemiology of Eye Diseases
SFA: saturated fat
T2D: type 2 diabetes
VLDL: very low-density lipoprotein

## Acknowledgements

All authors have no conflicts of interest.

CS, SN, TYW, C-YC, PJ, AL, MP, VS, EST, PW, RMvD and XS and conceived and designed research. AC and PW conducted experiments. CS, SN, TYW, C-YC, PJ, AL, MP, VS, EST, PW and RMvD contributed data. JYHS, YH, AC and XS analyzed data. JYHS, RMvD and XS wrote the manuscript and had primary responsibility for final content. All authors read and approved the manuscript.

The authors thank all investigators, staff members, and study participants for their contributions to all the participating studies. The MEC is supported by grants from the Singapore Ministry of Health, including National Medical Research Council Large Collaborative Grant (MOH-000271-00), National University of Singapore and National University Health System, Singapore. Metabolic profiling for MEC was supported by NUHS Summit Research Program (Metabolic Diseases) and performed by Nightingale Health Ltd. The SEED cohorts are supported by the National Medical Research Council (NMRC), Singapore (grants 0796/2003, 1176/2008, 1149/2008, STaR/0003/2008, 1249/2010, CG/SERI/2010, CIRG/1371/2013, CIRG/1417/2015, CIRG/1488/2018 and OFLCG/004a/2018), and Biomedical Research Council (BMRC), Singapore (08/1/35/19/550 and 09/1/35/19/616). VS was supported by the Finnish Foundation for Cardiovascular Research. JYHS was supported by the Singapore Ministry of Health’s National Medical Research Council Large Collaborative Grant (MOH-000271-00).

