## Supplemental Text for "Circulating metabolic biomarkers are consistently associated with incident type 2 diabetes in Asian and European populations – a metabolomics analysis in five prospective cohorts"

**Supplemental text: Cohort descriptions and ascertainment of type 2 diabetes**

*Multiethnic Cohort (MEC)*

The methods of MEC have been previously described (1). Briefly, the MEC was comprised of individuals who participated in previous population-based studies conducted in Singapore and additional enrollment to increase the numbers of ethnic minority groups. We conducted baseline assessments from 2004 until 2010 in 14,465 participants with a good representation of Chinese, Malay, and Indian ethnic groups, the three main ethnic groups in Singapore.

At recruitment, we collected information on socio-demographic and lifestyle factors through standardized interviewer-administered questionnaires. We assessed education level by asking participants their highest education level attained, from six options (‘no formal qualifications/lower primary’, ‘Primary (Primary School Leaving Examination)’, Secondary ('O'/'N' level)’, ‘Institute of Technical Education/National Technical Certificate’, ‘'A' level/ Polytechnic/ diploma’ and ‘University’). The questionnaire also ascertained medical history of the participants. Subsequently, we invited participants to a physical examination during which we conducted anthropometric and blood pressure measurements. Participants were instructed to remove their shoes before we measured their height on a portable stadiometer (SECA 200 series, Germany) in the Frankfurt Plane position. The weight of participants was measured on SECA digital scales (SECA digital scales (SECA 700 series, Germany) after participants removed all objects from pockets. BMI was then calculated by dividing the weight (kg) by the square of a participant’s height (m^2^). We collected two readings of systolic and diastolic blood pressure measurements using an automated digital monitor (Dinamap Carescape V100, General Electric) after the participants rested for 5 min. If the difference between the first two readings was >10 mmHg (for systolic blood pressure) or 5 mmHg (for diastolic blood pressure), we took a third set of reading for systolic and diastolic blood pressure. We used a sphygmomanometer (Accoson, UK) for the small minority of participants whose blood pressure exceeded the range of the automated digital monitor. Mean values of blood pressures were used in subsequent analyses. We also collected 8 to 12 h fasting blood samples from which clinical risk factors including fasting glucose and triglycerides were measured on the same day using enzymatic or other validated analytical (e.g. immunoassay, spectrophotometry) methods (1). Blood samples were analyzed at the biochemistry laboratory of the National University Hospital (April 2007 to October 2007 and November 2010 to August 2016) or the Singapore General Hospital biochemistry laboratory (October 2007 to November 2010).

We obtained ethics approval from SingHealth Centralised Institutional Review Board and the National University of Singapore Institutional Review Board.

*Singapore Epidemiology of Eye Diseases (SEED) cohorts*

The SEED cohorts consist of the Singapore Chinese Eye Study, Singapore Malay Eye Study and the Singapore Indian Eye Study which are population-based studies involving Chinese, Malay and Indian participants aged 40 to 80 years, conducted in Singapore from 2004 to 2011. Details of these cohorts have been published elsewhere (2, 3). Using an age-stratified sampling strategy, 6752 Chinese, 5600 Malay and 6350 ethnic Indian potential participants were selected from the Ministry of Home Affairs in Singapore. Potential participants who were deceased or terminally ill, or who were not living at the listed residential address were deemed ineligible. A total of 3353 Chinese, 3280 Malays and 3400 ethnic Indians consented to participate.

All participants were visited at home and were asked to complete a standardized interviewer-administered questionnaire that ascertained information on self-reported socio-demography (including age, sex and education level), lifestyle factors and medical history. The recruitment also consisted of a standardized examination procedure that included anthropometric and blood pressure measurements. During the examination, we measured height using a wall-mounted measuring tape and weight using a digital scale (SECA, model 782 2321009; Vogel & Halke, Germany). We then computed BMI by dividing the weight (kg) by the square of a participant’s height (m^2^). We measured systolic and diastolic blood pressure with a digital automatic blood pressure monitor (Dinamap model Pro Series DP110X-RW, 100V2; GE Medical Systems Information Technologies, Inc., USA) after participants were instructed to rest for 5 min. Two sets of readings were taken 5 min apart. We took a third measurement if the difference between the first two readings was >10 mmHg (for systolic blood pressure) or 5 mmHg (for diastolic blood pressure). The blood pressure of the participant was computed as the mean between the two closest readings. We also collected a 40 mL sample of non-fasting venous blood sample for biochemistry tests including glycosylated hemoglobin A1c (HbA1c). Biochemical measurements were performed by the National University Hospital Reference Laboratory on the same day as blood collection. Additional plasma (SEED Malays) and serum (SEED Chinese and SEED Indians) was subsequently stored at -80^◦^ C.

We obtained ethics approval from the Singapore Eye Research Institute (SERI) Institutional Review Board.

*FINRISK and Health 2000 cohorts*

FINRISK 2002, 2012 and Health 2000 are the Finnish population-based cohorts conducted by the Finnish Institute for Health and Welfare (4, 5). The FINRISK cohort studies were initiated with the goal of assessing cardiovascular risk and later also other chronic diseases risk in the Finnish population. The FINRISK health examination surveys were undertaken every five years beginning in 1972 in provinces of North Karelia, Northern Savo, North Ostrobothnia, Kainuu and five Finnish cities (Turku, Loimaa, Helsinki, Vantaa and Oulu). Each survey has added a new cohort into the FINRISK Study. The cohorts included in the present work were initiated in 2002 and 2012, in which 8775 and 5813 participants consented to join, respectively. Health 2000 was carried out during 2000 to 2001 in 80 regions across Finland (n = 8028). All cohorts involved newly selected randomly chosen participants aged 25 to 98 (25 to 74 in FINRISK, and 30 and over in Health 2000).

All surveys involved a self-administered questionnaire and a health examination. The questionnaire collected information on socio-demographic and lifestyle factors, and medical history. During the health examination, height was measured using a stadiometer to the nearest 0.1 cm after participants were asked to remove their shoes. Participants’ weight was measured with a beam-balanced scale to the nearest 0.1 kg without shoes and wearing only light clothing. For every participant, BMI was calculated as the weight (kg) divided by the square of height (m^2^). Venous blood samples were collected during health examination and sent to laboratories at the National Institute for Health and Welfare for analysis of biomarkers including fasting blood glucose (for Health 2000), HbA1c (for FINRISK 2002 and FINRISK 2012) and plasma lipids.

All study participants have been followed using nationwide electronic health registries, including national hospital discharge register, causes-of-death register, and drug reimbursement register. The Coordinating Ethical Committee of the Helsinki and Uusimaa Hospital District, Finland approved the cohort studies. Written informed consent was obtained from all participants.
